# An increase in aspartate aminotransferase levels can predict worsening disease severity in Japanese patients with COVID-19

**DOI:** 10.1101/2024.04.23.24306214

**Authors:** Kengo Matsumoto, Tsutomu Nishida, Satoru Okabe, Naohiro Sakamoto, Yoshifumi Fujii, Naoto Osugi, Aya Sugimoto, Dai Nakamatsu, Masashi Yamamoto, Koji Fukui, Osamu Morimura, Kinya Abe, Yukiyoshi Okauchi, Hiromi Iwahashi, Masami Inada

## Abstract

**Background:** The prognostic significance of liver dysfunction in coronavirus disease 2019 (COVID-19) patients remains unclear. This study aimed to investigate the association between liver function test results and severe disease progression in COVID-19 patients.

**Methods:** We conducted a retrospective study that included consecutive Japanese COVID-19 patients between February 2020 and May 2021. We identified the predictive variables for severe disease progression by utilizing established factors and multivariate logistic analysis. The Kaplan‒Meier method was used to estimate severe disease-free survival. Furthermore, we evaluated the hazard ratios (HRs) among three aspartate aminotransferase (AST) grades using Cox regression analysis: grade 1, AST < 30 U/L; grade 2, 30 U/L≤ AST < 60 U/L; and grade 3, AST >60 U/L.

**Results:** After exclusion, 604 symptomatic COVID-19 patients were enrolled during the study period, and 141 (23.3%) of them developed severe disease at a median of 2 days postadmission. The median hospital stay was 10 days, and 43 patients (7.1%) died during hospitalization. Multivariate regression analysis of the fourteen significant variables revealed that hypertension, decreased lymphocyte count, and elevated LDH, CRP, and AST levels (grade 2 and grade 3 relative to grade 1) were significant predictive variables. Severe disease-free survival times were significantly separated according to AST grade severity (HR: grade 2 to grade 1: 4.07 (95% CI: 2.06-8.03); HR: grade 3 to grade 1: 7.66 (95% CI: 3.89-15.1)).

**Conclusion:** AST levels at admission were an independent risk factor for severe disease in hospitalized Japanese patients with COVID-19.

## Introduction

In December 2019, Wuhan, China, experienced an outbreak of severe acute respiratory syndrome coronavirus 2 (SARS-CoV-2), which was subsequently named coronavirus disease 2019 (COVID-19) by the World Health Organization (WHO) in February 2020. The disease has spread rapidly, leading to a global pandemic that has persisted for three years to date. The first case of COVID-19 pneumonia in Japan was reported in Kanagawa Prefecture on January 15, 2020. The number of cases peaked in early April, confirming an epidemic (the first wave of the COVID-19 pandemic). Subsequently, the number of cases increased among individuals aged 20 to 30 years, reaching its peak in early August (the second wave). Outbreaks occurred in early January 2021 (the third wave), early May 2021 (the fourth wave, primarily caused by α strains), late August 2021 (the fifth wave, predominantly caused by δ strains), and January 2022 (the 6th wave, when Omicron strains rapidly replaced δ strains). Although the greatest number of patients were recorded in the seventh wave (caused by Omicron strains), the number of patients with severe disease requiring respiratory care was relatively small. As of July 21, 2022, in Japan, the Omicron BA.2 strain, a substrain of the B.1.1.529 strain, was believed to have been replaced by the BA.5 strain, which has become the dominant strain. [1]

Although COVID-19 has been reported to cause liver dysfunction in both Japanese and international studies, the pathogenesis, treatment, and prognostic significance of hepatic dysfunction in COVID-19 patients remain unclear. [2] Recent reports indicate that more than half of COVID-19 patients exhibit varying levels of liver dysfunction. Elevated aminotransferase levels have been reported in 14–58% of hospitalized patients with COVID-19. [3] While the degree of aminotransferase level increase is usually mild (<5 times the upper normal limit), severe acute hepatitis and high aminotransferase levels have been documented. However, comprehensive data on the clinical characteristics of liver enzyme elevation and liver failure in Japanese COVID-19 patients are lacking.

Although numerous risk factors for severe COVID-19 have been identified, the impact of liver function abnormalities has yet to be investigated owing to confounding factors and the limited number of reports in large Japanese patient cohorts. In this study, we aimed to retrospectively examine the associations between liver function test values and clinical characteristics in our cohort of Japanese COVID-19 patients who were diagnosed before the 6th wave and had a lower rate of severe disease.

## Patients and Methods

This was a retrospective single-center cohort study of consecutive Japanese patients who were diagnosed with COVID-19 through PCR or antigen methods and admitted to our hospital between February 2020 and July 2021. Osaka Prefecturure, the third most populous prefecture in Japan, established the Osaka Prefectural Inpatient Follow-up Center on March 30, 2020, to coordinate hospitalization based on patient symptoms and triaged patients with COVID-19 according to disease severity. This public hospital, located in Toyonaka in Osaka Prefecture, has a population of approximately 400,000 and is primarily designated a general hospital for treating patients with moderate to severe COVID-19. This has been in place since the first wave of the pandemic in Japan. [4-8]

We assessed the severity and treatment strategy for hospitalized COVID-19 patients based on guidance from the Japanese Ministry of Health, Labor, and Welfare available on the COVID-19 website, which has been available since March 7, 2020. The latest version (8.0) was released in July 2022 (in Japanese). [1] The severity of COVID-19 was categorized into four groups: mild disease without respiratory symptoms (SpO_2_ ≥96%); moderate I disease with breathing difficulties or pneumonia without respiratory failure (93%<SpO_2_<96%); moderate II disease requiring oxygen support (SpO_2_ ≤93%); and severe disease requiring intensive care unit (ICU) treatment or intubation. This Japanese severity classification is generally equivalent to the WHO severity classification for mild, moderate, severe, and critical disease [9], although there are some differences in the definitions. Treatment strategies have been updated with therapeutic advances. The therapeutic agents used in our hospital were hydroxychloroquine, favipiravir, ciclesonide, and dexamethasone from waves 1 to 3 and favipiravir, dexamethasone, heparin, and <u>remdesivir</u> from waves 4 to 5. We reviewed the clinical characteristics of COVID-19 patients with liver involvement by collecting electronic medical records from our hospital (MegaOak Online Imaging System, NEC, Japan). Clinical and laboratory data were collected from all patients at hospital admission. We prospectively used medical templates to assess all hospitalized COVID-19 patients at our infectious disease unit to avoid missing data. Patient height and body weight were obtained from medical interviews to reduce contact with patients. We conducted hepatitis B virus serology and hepatitis C virus antibody tests if they had not been previously conducted. We performed a portable chest radiograph for the initial evaluation of pulmonary complications at admission.

This study was conducted in accordance with the Declaration of Helsinki, and the institutional review board of Toyonaka Municipal Hospital approved this study (2022-10-3). The requirement for informed consent was waived by the same committee using the opt-out method on our hospital website.

In the context of our retrospective study, data were accessed on December 11, 2022, for research purposes.Throughout the research process, including during and after data collection, we had access to information that could potentially identify individual participants.All necessary ethical considerations and precautions were undertaken to maintain confidentiality and comply with applicable data protection regulations.

We investigated several predictive variables associated with the development of severe conditions, including age ≥ 65 years, BMI ≥30, smoking status, hypertension status, diabetes status, hyperlipidemia status, chronic kidney disease status, chronic lung disease status, solid cancer status, pregnancy status, lymphocyte count, lactate dehydrogenase (LDH) level, and C-reactive protein (CRP) level. We previously reported that the cutoff values for lymphocyte count, LDH levels, CRP levels, and estimated glomerular filtration rate (eGFR) were 980 count/μl, 309 IU/ml, 2.92 mg/dl, and 68 ml/min, respectively, as predictive risk factors in our hospital cohort. [4] In the present study, we estimated that cutoff values of 1000, 300, 3, and 70 ml/min (CKD) were predictive risk factors. The FIB-4 index, which evaluates liver fibrosis, was computed using the formula FIB-4 index = age × aspartate aminotransferase (AST)/platelet count × √ alanine aminotransferase (ALT). [10] We also assessed two liver function parameters, aspartate aminotransferase (AST) and alanine aminotransferase (ALT), and categorized patients into three groups at admission based on their AST and ALT levels: those with normal aminotransferase levels (grade 1, AST/ALT < 30 U/L), those with moderately elevated aminotransferase levels (grade 2, 30 U/L≤ AST/ALT < 60 U/L), and those with highly elevated aminotransferase levels (grade 3, 60 U/L< AST/ALT).

### Statistical analysis

Continuous variables are presented as medians and interquartile ranges (IQRs), and categorical variables are summarized as frequencies (%). We used Fisher’s exact test to evaluate differences in categorical variables and the Kruskal–Wallis test to compare differences in continuous variables among the three groups. We performed a univariate logistic analysis of the known risk factors for increased severity and stratified the patients into three groups according to their AST and ALT levels to identify significant risk factors. When both AST and ALT levels were identified as significant factors in the univariate analysis, we selected statistically significant factors with higher odds ratios due to confounding factors. We used logistic multivariate analysis with the extracted factors to examine the effects of liver dysfunction test values on illness severity. We estimated the severe disease-free survival time from admission using the Kaplan‒Meier method and evaluated hazard ratios (HRs) among the three AST groups using Cox regression analysis.

All calculated P values were two-tailed, with a P value < 0.05 considered to indicate statistical significance. We performed all the statistical analyses using JMP statistical software (ver. 16.20; SAS Institute Inc.).

## Results

During the study period, 748 COVID-19 patients were enrolled. Of these, 37 readmitted patients, 17 patients transferred from another hospital, 29 asymptomatic patients, 41 patients younger than 16 years, and 10 non-Japanese patients were excluded, as were 10 patients with insufficient data. Ultimately, 604 patients were included in the final analysis (**Figure 1**).

**Figure 1:**
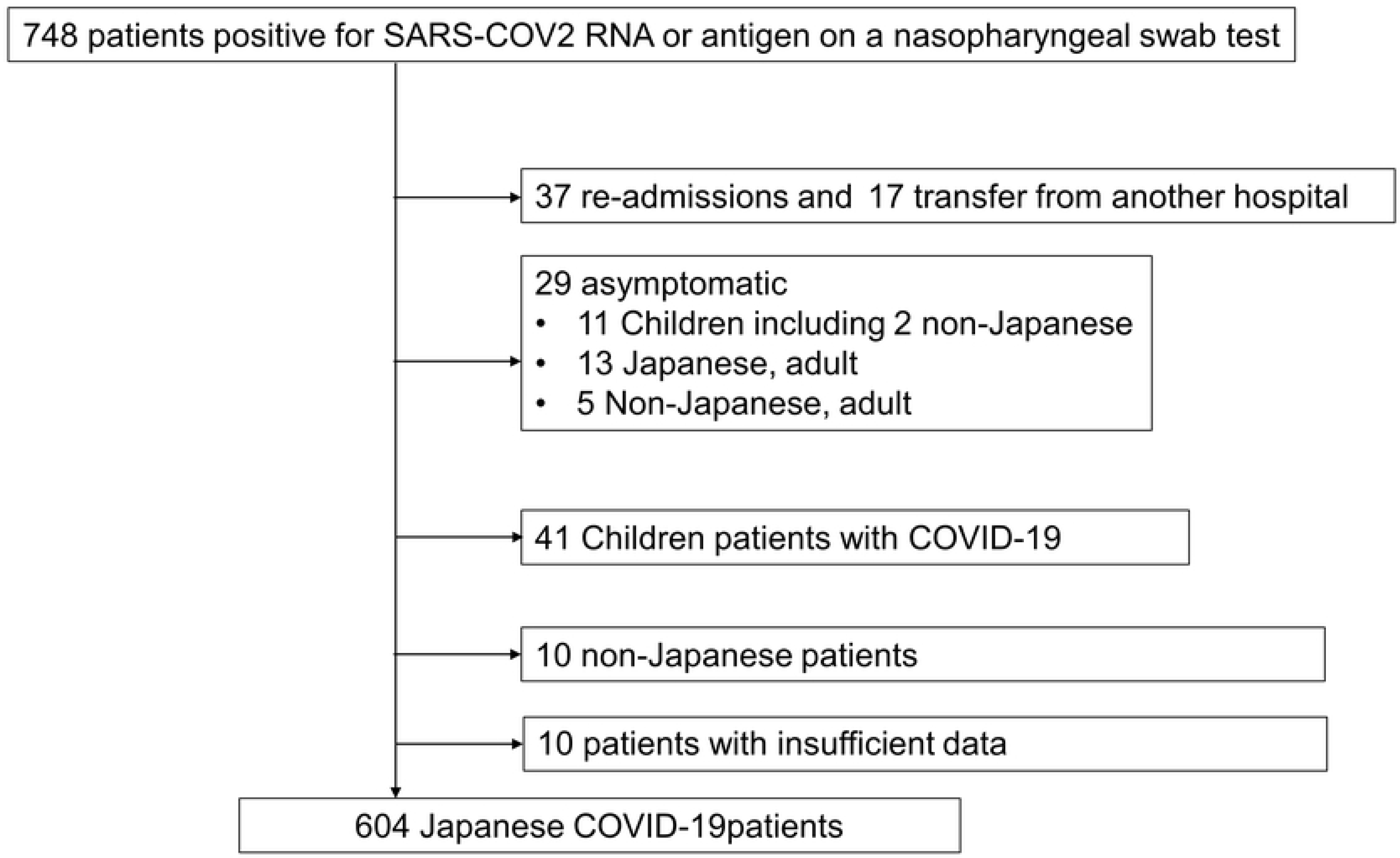
Flow chart of patient enrollment. COVID-19. We enrolled 748 COVID-19 patients during the study period, and 604 patients were included in the final analysis.

The clinical characteristics of the enrolled COVID-19 patients are summarized in **Table 1**. The median age was 62 years (IQR, 47-78), and 335 patients (55.5%) were men. The median BMI was 23.9 (IQR 21.0, 26.9), and 34.9% and 34.9% of the patients had a history of alcohol consumption and smoking, respectively. Other comorbidities included hypertension in 270 (44.1%) patients, cardiac disease in 103 (17.2%), diabetes in 168 (27.9%), hyperlipidemia in 133 (23.3%), and pregnancy in 23 (3.8%). Regarding medications, 148 (24.5%) patients were taking ACEis/ARBs, 190 (31.6%) were taking calcium blockers, and 110 (18.3%) were taking statins.

**Table 1.** Characteristics of symptomatic patients with confirmed COVID-19 on admission.

| Characteristics | Patients with COVID-19, n=604 |
| --- | --- |
| Age, median (IQR) | 62 (47, 78) |
| Men Sex, n (%) | 335 (55.5) |
| Body mass index, median (IQR) | 23.9 (21.0, 26.9) |
| Smoking history (Non/past/current) | 393/33/178 |
| Drinking history, yes (%) | 211 (34.9) |
| Days from the onset of symptoms to admission, median (IQR) | 6 (4, 9) |
| Laboratory data on admission |  |
| WBC, median (IQR) count/μl | 5100 (4050, 6800) |
| Lymphocyte, median (IQR) (count/μl) | 920 (672, 1240) |
| Neutrophilia, median (IQR) (count/μl) | 3699 (2663, 5252) |
| Hemoglobin, median (IQR) (g/dl) | 13.7 (12.3, 14.8) |
| Platelet count, median (IQR) ($10^9$/L) | 18.4 (14.7, 23.0) |
| AST, median (IQR) (IU/L) | 32 (23, 49) |
| ALT, median (IQR) (IU/L) | 24 (15, 40) |
| ALP, median (IQR) (IU/L) | 168 (74, 216) |
| $\gamma$ GTP, median (IQR) (IU/L) | 35 (18, 69) |
| T-bil, median (IQR) (mg/dl) | 0.55 (0.4, 0.71) |
| FIB-4 index, median (IQR) | 2.19 (1.32, 3.60) |
| Cr, median (IQR) (mg/dl) | 0.81 (0.63, 1.10) |
| BUN, median (IQR) (mg/dl) | 14 (11, 20) |
| LDH, median (IQR) (IU/L) | 262 (201, 356) |
| CRP, median (IQR) (mg/dl) | 3.6 (0.78, 7.9) |
| eGFR (mL/min/1.73m<sup>2</sup>), median (IQR) | 69.1 (50.7, 87.7) |
| HbA1c, median, median (IQR) (%) | 6 (5.7, 6.6) |
| Casual blood glucose, median (IQR) (mg/dL) | 115 (100, 140) |
| T-Chol, median (IQR) (mg/dL) | 164 (142, 189) |
| Comorbidities |  |
| Hypertension, n (%) | 270 (44.1) |
| Cardiovascular diseases, n (%) | 103 (17.2) |
| Chronic obstructive pulmonary disease, n (%) | 32 (5.4) |
| Asthma, n (%) | 49 (8.2) |
| Diabetes mellitus, n (%) | 168 (27.9) |
| Hyperlipidemia, n (%) | 133 (23.3) |
| Chronic kidney disease, n (%) | 71 (11.9) |
| Hemodialysis, n (%) | 36 (6.1) |
| Solid cancer, n (%) | 48 (8.0) |
| Pregnancy, n (%) | 23 (3.8) |
| Concomitant liver disease |  |
| HBs Ag positive, n (%) | 3 (0.53) |
| HCV Ab positive n (%) | 18 (3.2) |
| AIH or PBC, n (%) | 0 (0) |
| Use of medication for comorbidities |  |
| ACEi/ARB, n (%) | 148 (24.5) |
| Calcium blocker, n (%) | 190 (31.6) |
| Statin, n (%) | 110 (18.3) |
| PPI, n (%) | 133 (22.2) |

IQR: *Interquartile range*, AIH: autoimmune hepatitis, LDH: lactate dehydrogenase, CRP: C-reactive protein, AST: aspartate aminotransferase, Cr: creatinine, BUN: blood urea nitrogen, eGFR: estimated glomerular filtration rate, ARB: angiotensin receptor blocker, PPI: PBC: primary biliary cholangitis

The median time from onset to admission was 6 days (IQR: 4–9 days). On admission, 92.7% of the patients had fever, 48% had fatigue, 77.3% had respiratory symptoms, and 75.6% had pneumonia. Twenty-five patients (4.1%) were diagnosed with severe disease on admission, and 141 (23.3%) developed severe disease within a median of 2 days (IQR 1,5) of hospitalization. The median length of hospital stay was 10 days (IQR 7,15), and 43 (7.1%) patients died during hospitalization (**Table 2**).

**Table 2.** Initial presentation on admission, treatment, and clinical course.

|  |  |
| --- | --- |
| Initial presentation |  |
| Fever, n (%) | 559 (92.7) |
| Fatigue, n (%) | 206 (48) |
| Respiratory related symptom, n (%) | 468 (77.3) |
| Pneumonia, n (%) | 456 (75.6) |
| Headache, n (%) | 55 (12.3) |
| New loss of taste or smell, n (%) | 117 (23.2) |
| Days from onset of symptoms to admission, median (IQR) | 5 (3, 8) |
| Severity of COVID-19 on admission |  |
| Mild to moderate symptom, n (%) | 579 (95.9) |
| Severe symptom, n (%) | 25 (4.1) |
| Progression to severe disease, n (%) | 141 (23.3) |
| Treatment |  |
| Required oxygen, n (%) | 375 (62.1) |
| Medication for COVID-19 |  |
| Ciclesonide, n (%) | 156 (31.2) |
| Hydroxychloroquine, n (%) | 16 (2.6) |
| Favipiravir, n (%) | 291 (48.1) |
| Heparin, n (%) | 52 (8.6) |
| Remdesivir, n (%) | 35 (5.8) |
| Dexamethasone, n (%) | 305 (50.5) |
| Clinical course |  |
| Peak AST, median (IQR) (IU/L) | 41 (27,64) |
| Peak ALT, median (IQR) (IU/L) | 39 (21,73) |
| Length of hospital stay, median (IQR) (days) | 10 (7,15) |
| Time from admission to aggravation (IQR) (days) | 2 (1,5) |
| Required mechanical ventilatory support, n (%) | 125 (24.3) |
| Mortality, n (%) | 43 (7.1) |

IQR: *Interquartile range*,

### Liver function tests on admission

The median levels of AST and ALT at admission were 32 IU/ml (23, 49) and 24 IU/ml (15, 40), respectively. The peak AST and ALT levels during hospitalization were 41 IU/ml (27, 64) and 39 IU/ml (21, 73), respectively. The median alkaline phosphatase (ALP), γ-glutamyl transferase (GGT), and total bilirubin levels and FIB-4 index score at admission were 168 IU/ml (74, 216), 35 IU/ml (18, 69), 0.55 mg/dl (0.4, 0.71), and 2.19 (1.32, 3.60), respectively.

### Univariate logistic analysis and univariate Cox proportional hazards analysis of risk factors for progression to severe COVID-19

We conducted a univariate logistic analysis of the 13 established risk factors [1] and AST and ALT levels to evaluate their associations with progression to severe disease among 579 patients. Patients with severe disease at admission were excluded from the analysis. Older age (OR: 2.1, p=.0002), hypertension (OR: 2.94, p<.0001), diabetes mellitus (OR: 1.84, p=.0003), a decreased eGFR (OR: 2.38, p<.0001), a decreased lymphocyte count (OR: 3.03, p<.0001), an elevated LDH level (OR: 3.88, p<.0001), an elevated CRP level (OR: 3.29, p<.0001) and higher AST and ALT grades (ORs for Grade 3 to Grade 1 and Grade 2 to Grade 1: 5.13 and 2.36, P<.0001 and P=0.0002, respectively) were significantly associated with progression to severe disease (**Table 3**). Among these factors, AST grade had a significantly greater OR than did ALT grade; thus, AST grade was selected as a risk factor for subsequent multivariate regression analysis. Multivariate regression analysis of 14 variables (excluding ALT grade) revealed hypertension (OR: 2.24, P =.0026), a decreased lymphocyte count (OR: 2.72, P <.0001), an elevated LDH level (OR: 1.87, P =.002), an elevated CRP level (OR: 1.96, P=0.016) and an elevated AST level (ORs of 1.83 for Grade 2 and 3.35 for Grade 1, P=.0038, and P=. 0009) were significantly associated with progression to severe disease. Among the significant risk factors at admission, the AST >60 group had the highest OR for predicting progression to severe disease compared to the AST <30 group.

**Table 3.**
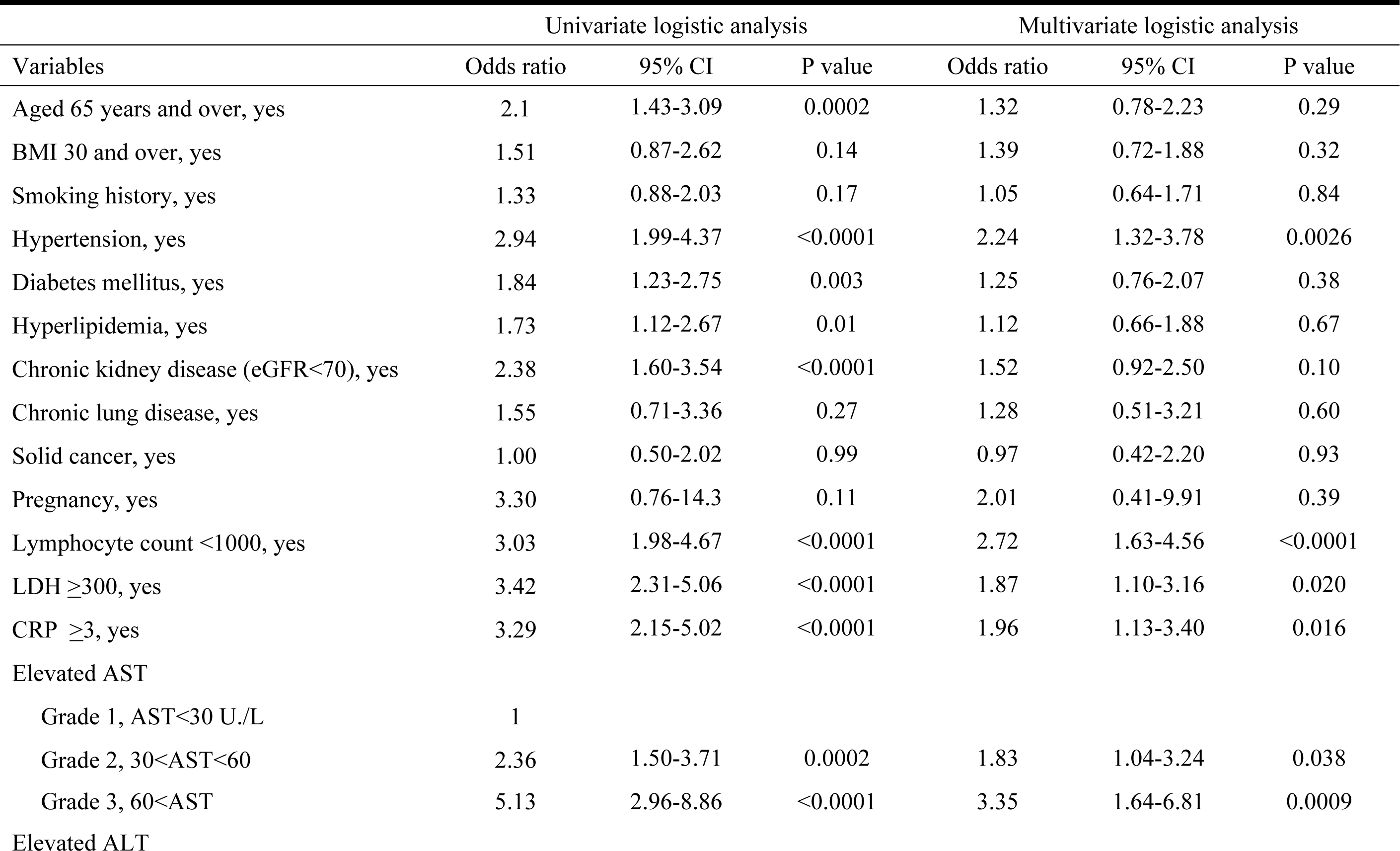
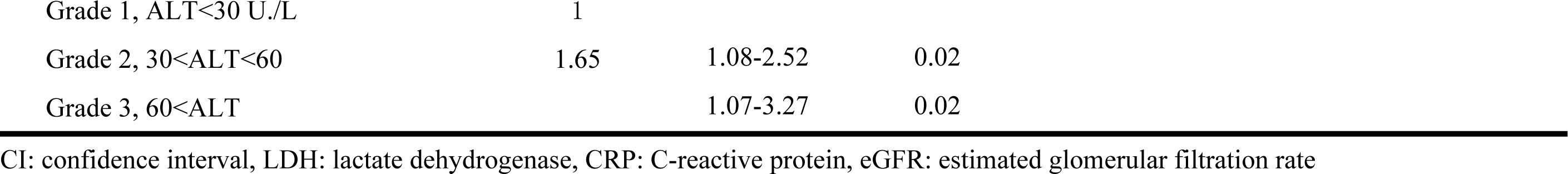
Univariate logistic analysis and multivariate regression analysis of risk factors for progression to critical COVID-19.

| Variables | Univariate logistic analysis |  |  | Multivariate logistic analysis |  |  |
| --- | --- | --- | --- | --- | --- | --- |
|  | Odds ratio | 95% CI | P value | Odds ratio | 95% CI | P value |
| Aged 65 years and over, yes | 2.1 | 1.43-3.09 | 0.0002 | 1.32 | 0.78-2.23 | 0.29 |
| BMI 30 and over, yes | 1.51 | 0.87-2.62 | 0.14 | 1.39 | 0.72-1.88 | 0.32 |
| Smoking history, yes | 1.33 | 0.88-2.03 | 0.17 | 1.05 | 0.64-1.71 | 0.84 |
| Hypertension, yes | 2.94 | 1.99-4.37 | <0.0001 | 2.24 | 1.32-3.78 | 0.0026 |
| Diabetes mellitus, yes | 1.84 | 1.23-2.75 | 0.003 | 1.25 | 0.76-2.07 | 0.38 |
| Hyperlipidemia, yes | 1.73 | 1.12-2.67 | 0.01 | 1.12 | 0.66-1.88 | 0.67 |
| Chronic kidney disease (eGFR<70), yes | 2.38 | 1.60-3.54 | <0.0001 | 1.52 | 0.92-2.50 | 0.10 |
| Chronic lung disease, yes | 1.55 | 0.71-3.36 | 0.27 | 1.28 | 0.51-3.21 | 0.60 |
| Solid cancer, yes | 1.00 | 0.50-2.02 | 0.99 | 0.97 | 0.42-2.20 | 0.93 |
| Pregnancy, yes | 3.30 | 0.76-14.3 | 0.11 | 2.01 | 0.41-9.91 | 0.39 |
| Lymphocyte count <1000, yes | 3.03 | 1.98-4.67 | <0.0001 | 2.72 | 1.63-4.56 | <0.0001 |
| LDH ≥300, yes | 3.42 | 2.31-5.06 | <0.0001 | 1.87 | 1.10-3.16 | 0.020 |
| CRP ≥3, yes | 3.29 | 2.15-5.02 | <0.0001 | 1.96 | 1.13-3.40 | 0.016 |
| Elevated AST |  |  |  |  |  |  |
| Grade 1, AST<30 U/L | 1 |  |  |  |  |  |
| Grade 2, 30<AST<60 | 2.36 | 1.50-3.71 | 0.0002 | 1.83 | 1.04-3.24 | 0.038 |
| Grade 3, 60<AST | 5.13 | 2.96-8.86 | <0.0001 | 3.35 | 1.64-6.81 | 0.0009 |
| Elevated ALT |  |  |  |  |  |  |

|  |  |  |  |
| --- | --- | --- | --- |
| Grade 1, ALT<30 U./L | 1 |  |  |
| Grade 2, 30<ALT<60 | 1.65 | 1.08-2.52 | 0.02 |
| Grade 3, 60<ALT |  | 1.07-3.27 | 0.02 |

CI: confidence interval, LDH: lactate dehydrogenase, CRP: C-reactive protein, eGFR: estimated glomerular filtration rate

### Clinical course of COVID-19 patients based on AST grade

**Table 4** compares the clinical characteristics of COVID-19 patients with AST levels of 1, 2 and 3 on admission. Among the 579 patients, 264 had grade 1 AST, 249 had grade 2 AST, and 91 had grade 3 AST on admission. Grade 1 restraint was significantly more prevalent in females and less prevalent in patients with a high BMI than in those with other BMIs. During the clinical course, 79 patients with grade 1 ASTs (30%) had worsened ASTs and were upgraded to higher ASTs (grade 2, n=54; grade 3, n=25), and 51 patients (20.5%) with grade 2 ASTs were upgraded to grade 3, whereas 383 patients with grades 1 and 2 (76%) did not have their ASTs changed (**Supplementary Figure 1**). The rates of progression to severe disease were 13.4% (35/259) for Grade 1 AST, 26.5% (66/236) for Grade 2 AST, and 44.0% (40/84) for Grade 3 AST (P<0.0001). The severe disease-free survival times in the three grade groups were significantly separated in parallel according to the AST severity (**Supplementary Figure 2**, HR of grade 2 to grade 1: 4.07 (95% CI: 2.06-8.03, P<.0001), HR of grade 3 to grade 1: 7.66 (95% CI: 3.89-15.1, P<.0001)).

**Table 4.**
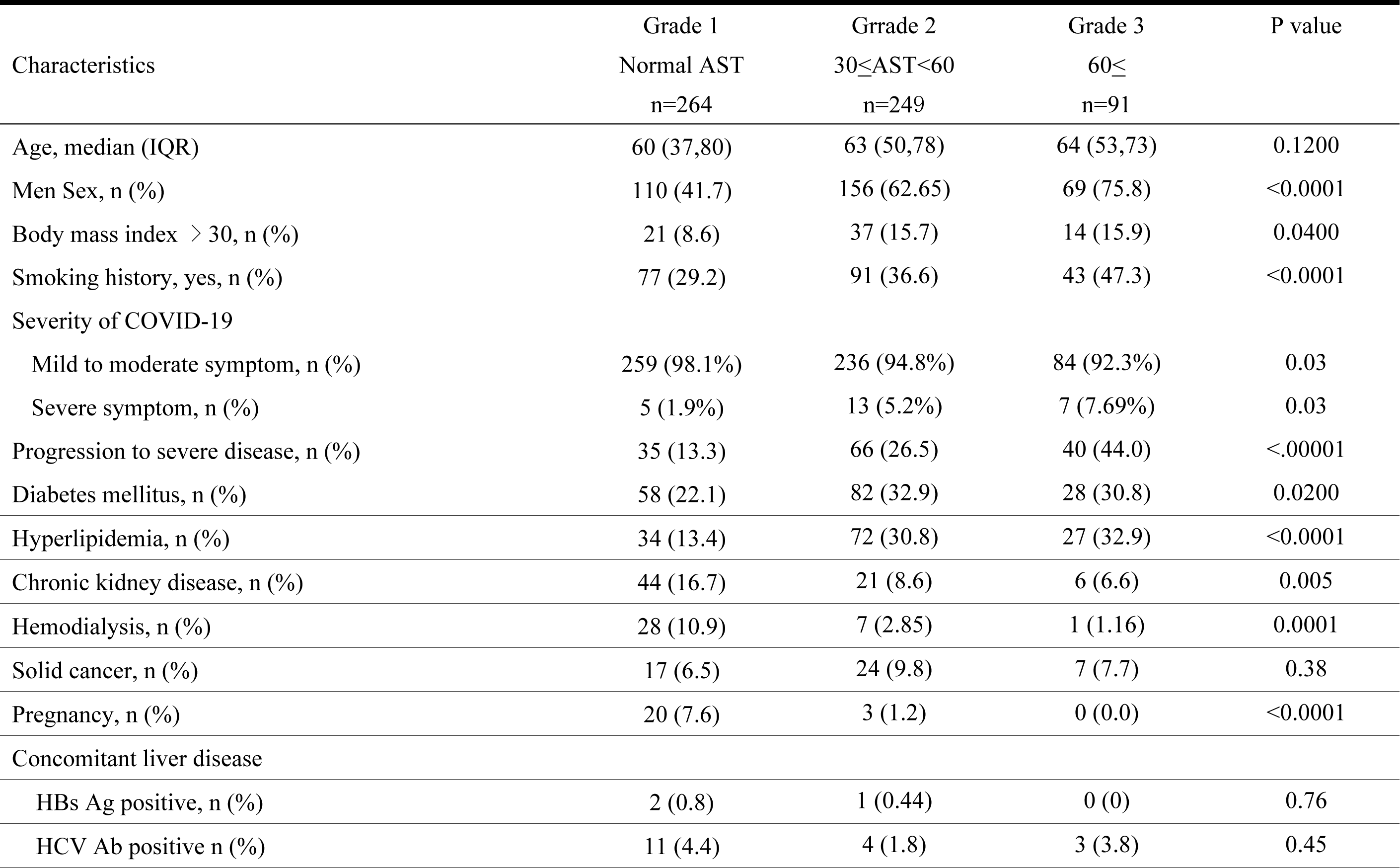
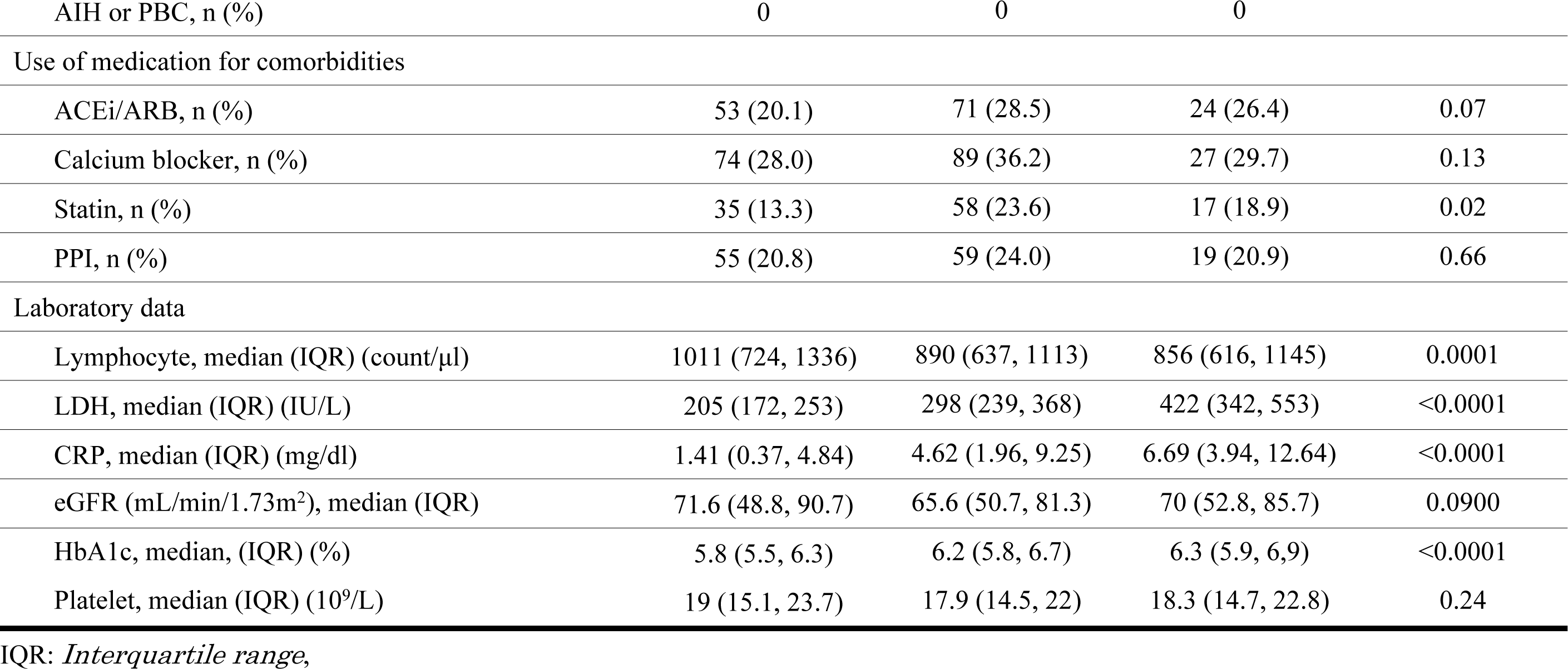
Comparison of three groups according to AST levels on admission.

### Patients with underlying liver disease

In the present study, we investigated patients with underlying liver disease. Overall, three patients were found to have positive hepatitis B surface (HBs) antigen tests, while 18 had positive hepatitis C (HCV) antibody tests. None of the patients had autoimmune hepatitis or primary biliary cholangitis during the study period, as indicated in **Table 5**. Notably, one out of three patients who had a positive HBs antigen test was found to be coinfected with HCV. Overall, 20 patients tested positive for viral hepatitis. Among these patients, six patients (30%) developed severe COVID-19. The median time from admission to progression to severe disease was three days (IQR: 0–3.5 days). The median AST and ALT levels were 29 IU/ml (IQR 23, 39) and 18 IU/ml (IQR 14, 32), respectively. Furthermore, the median peak AST and ALT levels during hospitalization were 40 IU/ml (IQR 26, 97) and 39 IU/ml (IQR 20, 71), respectively. The AST grade at the time of admission was 1 for 13 patients, 2 for four patients, and 3 for three patients. During the clinical course, six patients (30.0%) experienced an increase in the AST grade, but 14 patients did not experience any change in the AST grade.

**Table 5.** Clinical course and liver function tests in patients with underlying liver disease.

| Characteristics | Patients with liver disease, n=20 |
| --- | --- |
| HBs Ag positive, n | 3 |
| HCV infection (Current/preexisting), n | 9/9 |
| Age, median (IQR) | 70 (48.5,78) |
| Men Sex, n (%) | 13 (65) |
| Laboratory data on admission |  |
| AST, median (IQR) (IU/L) | 29 (23, 39) |
| AST levels on admission n (Grade 1/Grade 2/Grade 3) | 13/4/3 |
| ALT, median (IQR) (IU/L) | 18 (14, 32) |
| Platelet count, median (IQR) (10<sup>9</sup>/L) | 13.55 (11.83, 17.33) |
| Fib-4 index, median (IQR) | 2.81 (1.71, 4.38) |
| Clinical course |  |
| Peak AST, median (IQR) (IU/L) | 40 (26, 97) |
| Peak AST levels n (Grade 1/Grade 2/Grade 3) | 8/6/6 |
| Peak ALT, median (IQR) (IU/L) | 39 (20, 71) |
| Progression to severe disease, n (%) | 6 (30) |
| Time from admission to severe disease (IQR) (days) | 3 (0, 3.5) |
| Mortality, n (%) | 3 (15%) |

IQR: *Interquartile range*

## Discussion

Blood tests can provide important information on the condition of patients with COVID-19 and aid in predicting their prognosis. Therefore, blood tests are recommended for hospitalized COVID-19 patients who are at risk for severe disease or those with moderate or severe disease. Several studies on biomarkers for disease progression that assist in determining illness severity and predicting patient prognosis have been conducted in Japan and internationally. The application of these markers is expected to enhance the quality of medical care and optimize the utilization of medical resources. However, the impact of liver function tests has not been adequately explored, and there is a paucity of research studies conducted on large cohorts of Japanese patients. In this study, multivariate analysis incorporating established risk factors revealed that elevated AST levels at admission were an independent and significant risk factor for severe disease in hospitalized Japanese COVID-19 patients. Furthermore, of the COVID-19 patients who presented with AST levels of 60 IU/ml or higher on admission, 44.0% progressed to severe disease, which was 1.8 times greater than the rate in the entire cohort.

Regarding the impact of underlying liver disease, no significant findings were obtained due to the low number of patients with viral hepatitis or autoimmune diseases in this cohort. Owing to the constraints of minimizing contact with patients, abdominal ultrasound (US) was not performed; thus, we were unable to verify previous data or identify patients with alcoholic liver disease or nonalcoholic fatty liver disease [11]. Although a reduction in the PLT is indicative of the severity of liver fibrosis [12], it was challenging to determine whether the patients had underlying liver disease based on the PLT in this study, as it has been reported that PLT decreases in patients with COVID-19 infection due to inflammation and intravascular coagulation disorders. [13]

The liver may be vulnerable to SARS-CoV-2 infection due to the presence of angiotensin-converting enzyme 2 (ACE2) receptors on biliary and hepatic epithelial cells. [14] The virus enters and harms target organs by binding to ACE2 receptors. [15,16] Autopsy results have confirmed the presence of viral RNA in liver tissue [17], indicating the possibility of direct hepatocellular damage caused by SARS-CoV-2. In our study, 97 patients received angiotensin receptor blockers (ARBs), 9 received angiotensin-converting enzyme (ACE) inhibitors, and 45% had hypertension. Among the 105 patients who were treated with ARBs and/or ACE inhibitors, 35.2% (37/105) progressed to severe disease, which was significantly greater than the percentage of patients (20.5%; 82/401) who did not receive these medications (P=0.00027, data not shown). In addition, AST severity was significantly greater in ARB and/or ACE inhibitor users than in nonusers (grade 1 in 36.5% of users and 50.1% of nonusers, grade 2 in 48.2% of users and 38.0% of nonusers, grade 3 in 15.5% of users and 11.9% of nonusers; P=0.0478; data not shown). Our findings suggested that ARB/ACE inhibitor use may upregulate ACE2 receptor expression in biliary and hepatic epithelial cells, which could lead to liver dysfunction in ARB/ACE inhibitor users. However, the impact of hypertension as a confounding factor could not be determined. Furthermore, liver dysfunction in COVID-19 patients may also be attributed to inflammation, cytokine storms, the use of therapeutic drugs and hypoxemia associated with respiratory failure.

This study described the first to fifth waves of the COVID-19 outbreak in Japan, during which different mutant strains were prevalent, leading to varying infectivity and symptoms. Since the number of available drugs increased from the fourth period, we classified patients into two groups: those in the earlier period up to the third period and those in the latter period after the fourth period. We investigated differences in the background and frequency of liver injury between these two groups. The frequency of liver injury at presentation was greater in the latter period (Grade 1 (48.8%, 35.6%), Grade 2 (39.9%, 43.4%), and Grade 3 (11.3%, 21.0%)). However, the frequency of AST level upgrades was not significantly different between the two groups (21.8%, 21.0%, P=0.81). Although there was no significant difference in the severity of admission between the two groups, the frequency of progression to severe disease was significantly greater in the latter group (20.0%, 28.8% P=0.013) **(Supplementary Table 1**). Although it was challenging to assess this due to differences in hospitalization criteria during each wave, it was suggested that the variation in viral strains might have an impact on liver damage.

With the spread of COVID-19, existing drugs have been repurposed, and several treatments have been approved, but liver damage has been reported with COVID-19 treatments. [18-22] Such treatments have also been reported to induce liver injury. [23]. At our hospital, chloroquine, favipiravir, ciclesonide, and dexamethasone were used from the first to the third wave, and favipiravir, decadron, heparin, and remdesivir were used from the fourth wave to the fifth wave. The impact of each drug on liver dysfunction was studied. Univariate logistic analysis of the six drugs, disease progression to severe COVID-19, and worsening AST levels during hospitalization indicated that the use of favipiravir, ciclesonide, and remdesivir was a significant factor in exacerbating COVID-19. Hepatic injury exacerbation after hospitalization was influenced by drug use as well as by worsening COVID-19 symptoms; however, drawing conclusions is challenging due to several confounding factors (**Supplementary Table 2**).

The present study has several limitations. First, it was a retrospective, single-center study. Second, the search for past liver function abnormalities was limited, as the patients were examined in isolation with minimal contact, leading to potentially inadequate evaluation of some patients with abnormal liver function. Thus, the study could not determine whether the presence or absence of background liver disease impacted the course of the disease. Third, the differences in virus strains and the involvement of the drugs used have not been adequately investigated. The relationship between drugs and liver injury was also examined; however, the backgrounds of patients treated with each drug and those who did not differ in terms of the decision to administer drugs were based on the symptoms of COVID-19, making it challenging to draw conclusions regarding drug effects.

In conclusion, our study demonstrated that AST levels at admission were a significant independent risk factor for severe disease in hospitalized Japanese patients with COVID-19.

## Data Availability

The data that support the findings of this study are available on request from the corresponding author Kengo Matsumoto. The data are not publicly available due to restrictions (e.g., they contain information that could compromise the privacy of the research participants).

## Acknowledgments

N/A

## Figure legends

**Supplementary Figure 1:** AST levels on admission and peak AST levels. During the clinical course, 79 patients with grade 1 ASTs were upgraded to a higher AST grade, and 51 patients with grade 2 ASTs were upgraded to grade 3, although the AST grade did not change in 383 patients with grades 1 and 2.

**Supplementary Figure 2:** The severe disease-free survival times in the three grade groups. The three groups were significantly separated in parallel according to the severity of AST.

